# BENEFIT-RISK ASSESSMENT OF COVID-19 VACCINE, MRNA (MRNA-1273) FOR MALES AGE 18-64 YEARS

**DOI:** 10.1101/2022.12.02.22283050

**Authors:** Osman N. Yogurtcu, Patrick R. Funk, Richard A. Forshee, Steven A. Anderson, Peter W. Marks, Hong Yang

## Abstract

Since authorization of the Moderna mRNA COVID-19 Vaccine, real-world evidence has indicated its effectiveness in preventing COVID-19 cases. However, increased cases of mRNA vaccine-associated myocarditis/pericarditis have been reported, predominantly in young adults and adolescents. The Food and Drug Administration conducted benefit-risk assessment to inform review of the Biologics License Application for use of the Moderna vaccine among individuals ages 18 years and older. We modeled benefit-risk per million individuals who receive two complete doses of the vaccine. Benefit endpoints were vaccine-preventable COVID-19 cases, hospitalizations, intensive care unit (ICU) admissions, and deaths. The risk endpoints were vaccine-related myocarditis/pericarditis cases, hospitalizations, ICU admissions and deaths. The analysis was conducted on the age-stratified male population, due to data signals and previous work showing males to be the main risk group. We constructed six scenarios to evaluate the impact of uncertainty associated with pandemic dynamics, vaccine effectiveness (VE) against novel variants, and rates of vaccine-associated myocarditis/pericarditis cases on the model results. For our most likely scenario, we assumed the US COVID-19 incidence was for the week of December 25, 2021, and a VE of 30% against cases and 72% against hospitalization with the Omicron-dominant strain. Our source for estimating vaccine-attributable myocarditis/pericarditis rates was FDA’s CBER Biologics Effectiveness and Safety (BEST) System databases. Overall, our results supported the conclusion that the benefits of the vaccine outweigh its risks. Remarkably, we predicted vaccinating one million 18-25 year-old males would prevent 82,484 cases, 4,766 hospitalizations, 1,144 ICU admissions, and 51 deaths due to COVID-19, comparing to 128 vaccine-attributable myocarditis/pericarditis cases, 110 hospitalizations, zero ICU admissions, and zero deaths. Uncertainties in the pandemic trajectory, effectiveness of vaccine against novel variants, and vaccine-attributable myocarditis/pericarditis rate are important limitations of our analysis. Also, the model does not evaluate potential long-term adverse effects due to either COVID-19 or vaccine-attributable myocarditis/pericarditis.

## 1. Introduction

The Moderna COVID-19 mRNA (mRNA-1273) vaccine has been available for use in persons 18 years of age and older in the United States under Emergency Use Authorization (EUA) and was recently licensed in January 2022. Since authorization of mRNA COVID-19 vaccines (Pfizer-BioNTech and Moderna) in December of 2020, real-world evidence from the pre-Omicron period has indicated the vaccines effectively prevent COVID-19 cases, hospitalizations, and deaths. However, cases of myocarditis and pericarditis associated with mRNA COVID-19 vaccines have been reported in the United States (US), especially in adolescents (for whom the Pfizer-BioNTech vaccine is authorized) and young adult males^1-3^ through the Biologics Effectiveness and Safety (BEST) system which is an active post-market surveillance system at the US Food and Drug Administration’s (FDA) Center for Biologics Evaluation and Research (CBER). CBER follows FDA’s structured framework^16^ when making decisions on product licensure. FDA previously conducted and published a benefit-risk assessment for use of COMIRNATY among ages 16 years and older^6^. In this paper, to advance the transparency regarding FDA’s decision-making process, we report its benefit-risk (B-R) assessment (using principles similar to FDA^6^ and CDC’s^4^ earlier assessment for a different vaccine) to inform regulatory decisions related to the Biologics License Application (BLA) for use of the Moderna vaccine among individuals 18 years of age and older.

The regulatory question addressed by our analyses is whether the benefits of vaccination outweigh the risks for the target population, considering the uncertainties of the evolving pandemic (changes in disease incidence and emergence of new variants) and the risk of myocarditis/pericarditis after vaccination, predominantly among young males, identified by post-authorization safety surveillance. Our assessment used the structured benefit-risk framework (BRF) including four key dimensions - Analysis of Condition, Current Treatment Options, Benefits, and Risks and Risk Management^5^. We assessed the benefits and risks per million males ages 18-64 years who are vaccinated with two complete doses of the Moderna vaccine. The metric is fixed at per one million individuals with two complete primary doses of vaccination) to compare benefits and risks consistently among the groups without knowing how vaccine uptake may vary. We estimated COVID-19 cases, hospitalizations, ICU admissions, and deaths prevented by the vaccination (benefits) and myocarditis/pericarditis cases, related hospitalizations, ICU admissions, and deaths attributable to the vaccine (risks). We chose these endpoints as they are the most clear and measurable endpoints for benefits and risks that have the greatest public health significance. Modeling was not conducted for females and individuals 65 years of age and older due to previous work showing a relative lower risk^6^ among these groups, resulting in too few cases of myocarditis/pericarditis after vaccination in these groups to reliably estimate myocarditis/pericarditis rates. These groups are also expected to have more favorable benefit-risk compared to males age 18-64 years based on available evidence.

## 2. Methods

### 2.1. Model Overview

We used similar methodology as presented in our earlier paper^6^. Details of our calculation may be seen in our Microsoft Excel model file in the Supplementary. We assessed the benefits and risks per million male population stratified by age: 18-25, 26-35, 36-45, 46-55 and 56-64 years since data indicated an age-dependent risk of myocarditis and pericarditis post vaccination. Those age groups aligned with the data collection stratification for COVID-19 outcomes. The model benefit endpoints were vaccine-preventable COVID-19 cases, hospitalizations, intensive care unit (ICU) admissions, and deaths. The model risk endpoints were vaccine related myocarditis/pericarditis cases, hospitalizations, ICU admissions, and deaths (Figure 1). Key model inputs include duration of vaccine protection, VE against COVID-19 cases and hospitalizations, age-specific COVID-19 case and hospitalization incidence rates, age-specific vaccine-attributable myocarditis case rate, hospitalizations, ICU admissions, and death rate.

**Figure 1.**
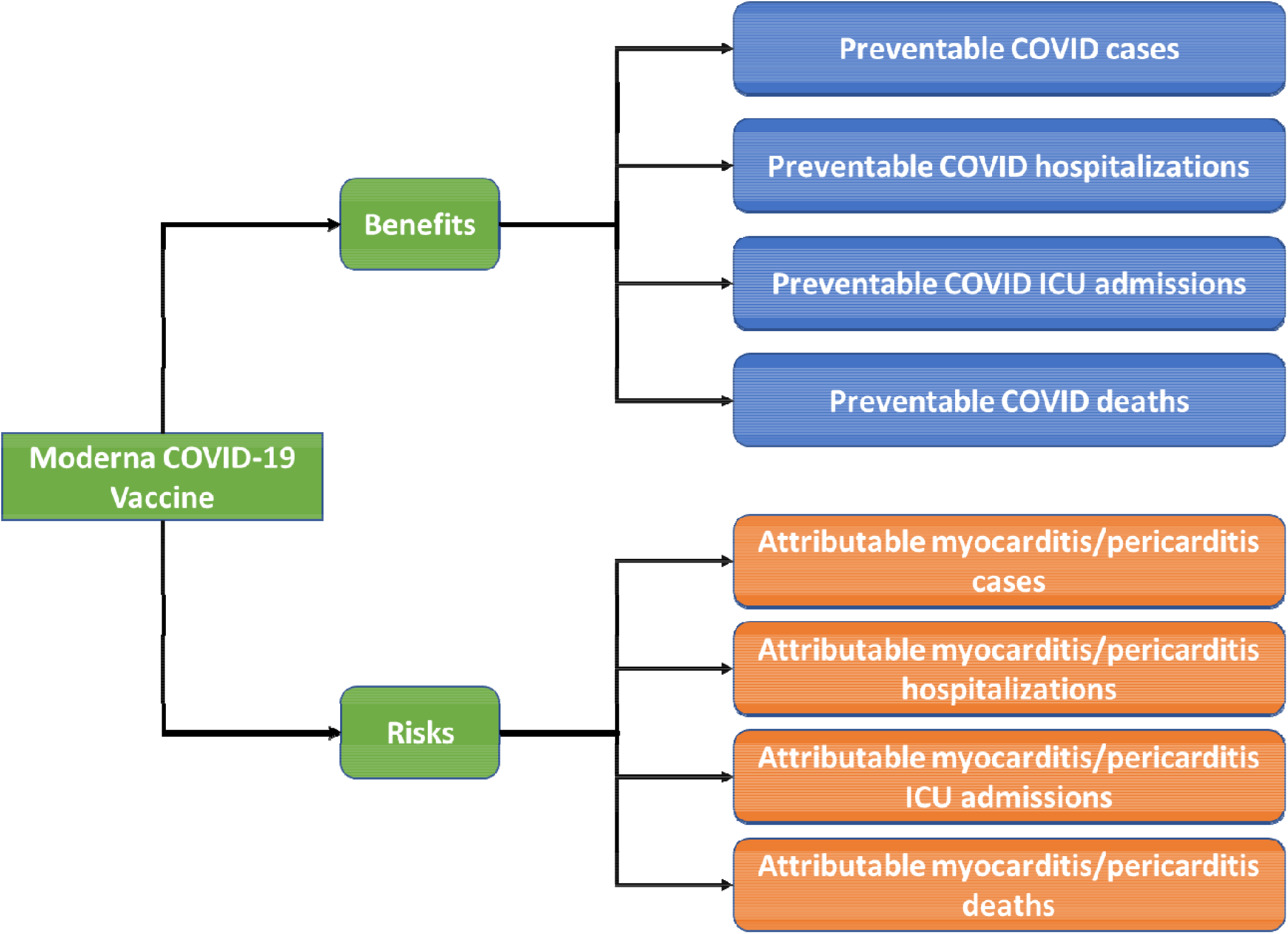
Benefits-risks value tree. Description: A tree from left to right pointing out the benefits and risks of Moderna CVmRNA.

Our model produced benefit-risk outcomes for six different scenarios (Table 1) as a sensitivity analysis of the uncertainties related to three major model inputs, COVID-19 incidence rate (Scenarios 1, 2, and 3), VE (Scenarios 1 and 4), and vaccine-attributable myocarditis/pericarditis rate (Scenarios 1, 5, and 6). We decide to use Scenario 1 as a base scenario since it represents the most-up-to date COVID-19 incidence at the time of this analysis. For the remaining scenarios, only one of the three major model inputs from Scenario 1 is modified at a time (summarized in Table 1). Model predictions are provided in Table 2. Visual benefit-risk plot for age groups 18-25, 26-35, and 36-45 for Scenario 1 is shown on Figure 2.

**Table 1.** Six model scenarios with varying COVID-19 incidence rates, vaccine effectiveness against cases and hospitalization, and myocarditis/pericarditis rates.

|  | COVID-19 Incidence | Vaccine Effectiveness | Excess Myocarditis Risk |
| --- | --- | --- | --- |
| Scenario 1 | As of December, 2021 | Omicron dominant:<br>30% against cases<br>72% against hospitalization<br>72% against death | Mean of BEST meta-analysis |
| Scenario 2 | Average COVID-19 pandemic incidence in 2021 | Same as Scenario 1 | Same as Scenario 1 |
| Scenario 3 | Lowest COVID-19 pandemic incidence (June 5, 2021) | Same as Scenario 1 | Same as Scenario 1 |
| Scenario 4 | Same as Scenario 1 | Delta dominant:<br>80% against cases<br>90% against hospitalization<br>90% against death | Same as Scenario 1 |
| Scenario 5 | Same as Scenario 1 | Same as Scenario 1 | 2.5<sup>th</sup> percentile of BEST meta-analysis |
| Scenario 6 | Same as Scenario 1 | Same as Scenario 1 | 97.5<sup>th</sup> percentile of BEST meta-analysis |

**Table 2.** Benefit-risk outcomes per million males vaccinated with 2 primary series doses of Moderna CVmRNA under each of six scenarios described in Table 1.

|  | Ages | BENEFITS |  |  |  | RISKS |  |  |  |
| --- | --- | --- | --- | --- | --- | --- | --- | --- | --- |
|  |  | COVID Cases | COVID Hospitalizations | COVID ICUs | COVID Deaths | Myo/Peri Cases | Myo/Peri Hospitalizations | Myo/Peri ICUs | Myo/Peri Deaths |
| Scenario 1 | 18-25yo | 82,484 | 4,766 | 1,144 | 51 | 128 | 110 | 0 | 0 |
|  | 26-35yo | 87,557 | 4,766 | 1,220 | 247 | 32 | 26 | 0 | 0 |
|  | 36-45yo | 90,939 | 4,766 | 1,258 | 378 | 23 | 18 | 0 | 0 |
|  | 46-55yo | 77,624 | 10,921 | 3,200 | 1,423 | 13 | 10 | 0 | 0 |
|  | 56-64yo | 68,747 | 15,025 | 4,928 | 2,120 | 10 | 7 | 0 | 0 |
| Scenario 2 | 18-25yo | 26,705 | 2,088 | 501 | 48 | 128 | 110 | 0 | 0 |
|  | 26-35yo | 29,718 | 2,088 | 535 | 235 | 32 | 26 | 0 | 0 |
|  | 36-45yo | 31,727 | 2,088 | 551 | 359 | 23 | 18 | 0 | 0 |
|  | 46-55yo | 29,174 | 4,607 | 1,350 | 1,087 | 13 | 10 | 0 | 0 |
|  | 56-64yo | 27,472 | 6,286 | 2,062 | 1,573 | 10 | 7 | 0 | 0 |
| Scenario 3 | 18-25yo | 3,903 | 635 | 152 | 7 | 128 | 110 | 0 | 0 |
|  | 26-35yo | 4,307 | 635 | 163 | 33 | 32 | 26 | 0 | 0 |
|  | 36-45yo | 4,576 | 635 | 168 | 50 | 23 | 18 | 0 | 0 |
|  | 46-55yo | 4,311 | 1,127 | 330 | 158 | 13 | 10 | 0 | 0 |
|  | 56-64yo | 4,134 | 1,456 | 477 | 230 | 10 | 7 | 0 | 0 |
| Scenario 4 | 18-25yo | 219,958 | 5,957 | 1,430 | 63 | 128 | 110 | 0 | 0 |
|  | 26-35yo | 233,486 | 5,957 | 1,525 | 309 | 32 | 26 | 0 | 0 |
|  | 36-45yo | 242,504 | 5,957 | 1,573 | 472 | 23 | 18 | 0 | 0 |
|  | 46-55yo | 206,998 | 13,652 | 4,000 | 1,779 | 13 | 10 | 0 | 0 |
|  | 56-64yo | 183,326 | 18,781 | 6,160 | 2,650 | 10 | 7 | 0 | 0 |
| Scenario 5 | 18-25yo | 82,484 | 4,766 | 1,144 | 51 | 68 | 58 | 0 | 0 |
|  | 26-35yo | 87,557 | 4,766 | 1,220 | 247 | 9 | 7 | 0 | 0 |
|  | 36-45yo | 90,939 | 4,766 | 1,258 | 378 | 11 | 9 | 0 | 0 |
|  | 46-55yo | 77,624 | 10,921 | 3,200 | 1,423 | 7 | 5 | 0 | 0 |
|  | 56-64yo | 68,747 | 15,025 | 4,928 | 2,120 | 4 | 3 | 0 | 0 |
| Scenario 6 | 18-25yo | 82,484 | 4,766 | 1,144 | 51 | 241 | 207 | 0 | 0 |
|  | 26-35yo | 87,557 | 4,766 | 1,220 | 247 | 119 | 97 | 0 | 0 |
|  | 36-45yo | 90,939 | 4,766 | 1,258 | 378 | 47 | 36 | 0 | 0 |
|  | 46-55yo | 77,624 | 10,921 | 3,200 | 1,423 | 23 | 18 | 0 | 0 |
|  | 56-64yo | 68,747 | 15,025 | 4,928 | 2,120 | 23 | 18 | 0 | 0 |

**Figure 2.**
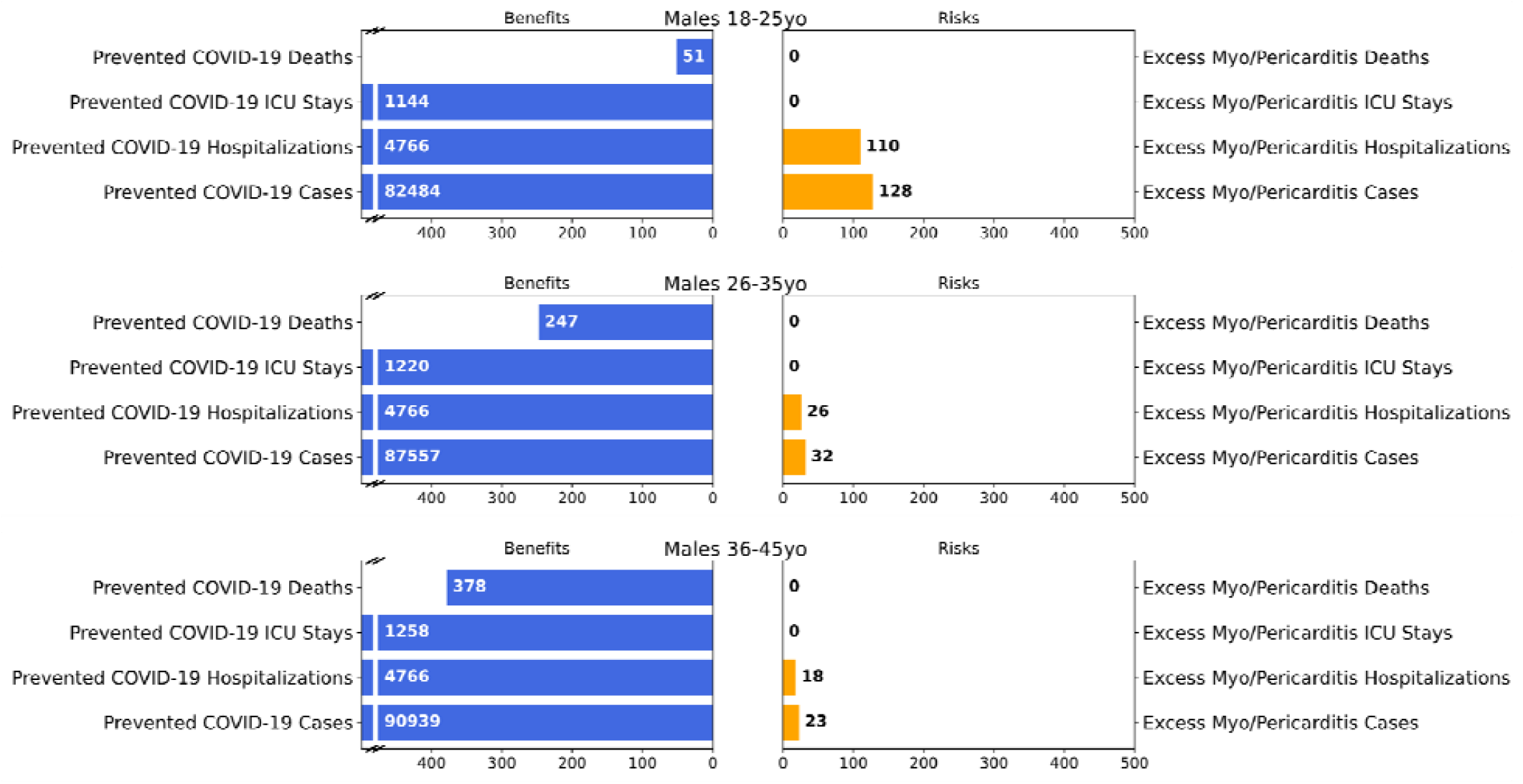
Benefit-risk outcomes per million males in age groups within 18-45 years for scenario 1 Description: Six bar graphs examining benefits (e.g., prevented deaths) and risks (e.g., excess deaths)

### 2.2. Benefits

#### 2.2.1. Data and assumptions

##### 2.2.1.1. Duration of vaccine protection

We estimated protection over a 5-month period after completion of the 2-dose primary series, since a 5-month interval between completion of the primary series and booster dose is authorized by FDA and recommended by the Centers for Disease Control and Prevention (CDC). For simplicity, the model did not account for benefits of partial vaccination and assumes a constant VE during the 5-month period post second dose. We did not factor in protection form potential exposure or infection with COVID-19 among unvaccinated individuals into our analysis since the degree of this protection and its time-dependency were not well understood at the time of analysis.

##### 2.2.1.2. Incidences of COVID-19 case, hospitalization, ICU, and death

The COVID-19 case and hospitalization incidence rates were assumed to remain constant over the assessment period (within 5 months post-second dose). For Scenarios 1, 4, 5, and 6, the crude incidence rates of COVID-19 cases and deaths in the unvaccinated population from the week of December 25, 2021, were obtained from the COVID Data Tracker for all male age groups^8^. The hospitalization rates for these scenarios were for the week of December 25, 2021, collected by COVID-NET^7^. The percentage of hospitalizations with ICU admission was estimated based on the cumulative rates of ICU admissions for each male age group from March 2020 to October 2021 provided by the CDC (collected by COVID-NET). We used the average 2021 US COVID-19 incidence rate and the lowest US COVID-19 incidence rate (June 2021) in Scenarios 2 and 3, respectively. All the incidence-related model inputs are summarized in Table 3.

**Table 3.** End of December 2021 US COVID incidences in the unvaccinated population listed by age groups (Male population only) and percentage of hospitalized going to ICU. Source: 1- COVID Data Tracker, 2-COVID-NET.

| **Age group** | **Daily COVID-19 cases/100k persons<sup>1</sup>** | **Daily Hospitalizations/ 100k persons<sup>2</sup>** | **Daily deaths/100k persons<sup>1</sup>** | **Percent of hospitalized going to ICU<sup>2</sup>** |
| --- | --- | --- | --- | --- |
| 18-25 | 180.89 | 4.35 | 0.05 | 24.0 |
| 26-35 | 192.01 | 4.35 | 0.23 | 25.6 |
| 36-45 | 199.43 | 4.35 | 0.34 | 26.4 |
| 46-55 | 170.23 | 9.98 | 1.30 | 29.3 |
| 56-64 | 150.76 | 13.73 | 1.94 | 32.8 |

##### 2.2.1.3. Vaccine effectiveness

We assumed Omicron as the dominant strain in Scenarios 1, 2, 3, 5, and 6 and assumed averages of 30% VE against COVID-19 cases and 72% VE against COVID-19 hospitalizations during the 5-month period post second dose. The data from a UK surveillance report were used to derive these VEs for Omicron^9^. For Scenario 4 we assume Delta as the dominant strain and used the US averages of 80% VE against cases and 90% VE against hospitalizations^10^. Many studies conducted in the US and other countries during the Delta-dominant period showed consistently high VE of the Moderna vaccine against both COVID-19 cases and related hospitalization^10,11^. For all scenarios, we assumed the vaccine effectiveness against death due to COVID-19 would be equal to the vaccine effectiveness against hospitalizations due to COVID-19.

### 2.3. Risks

#### 2.3.1. Calculation of risks

Our benefit-risk model had four risk endpoints (Figure 1): vaccine-attributable myocarditis/pericarditis cases, hospitalizations, ICU admissions, and deaths. Estimates of vaccine-attributable cases of myocarditis/pericarditis (per 1 million person-days with a risk window of 7 days post vaccination) were based on a meta-analysis of four health claims databases in BEST, which combined data from four data partners (DP) starting from December 10, 2020 (Table 4). Data cutoff dates for the sources were as follows: DP1 (August 21, 2021), DP2 (July 10, 2021), DP3 (July 31, 2021), and DP4 (June 30, 2021). The age-specific vaccine-attributable incidences of myocarditis/pericarditis were calculated for dose 2 and used as the model input for the risk of individuals with complete two dose vaccination. The risk post dose 1 was ignored since CDC has suggested people who have myocarditis after dose 1 to have a precaution to receive dose 2, and the majority the myocarditis cases were reported post dose 2. For Scenarios 1, 2, 3, and 4 we used the mean meta-analysis predicted myocarditis/pericarditis rate for each male age group, while for Scenario 5 we use the 2.5^th^ percentile rate and for Scenario 6 we used the 97.5^th^ percentile rate. We used Equation 1 to calculate total myocarditis/pericarditis (M_Pred_) per one million individuals who completed primary vaccination series (two doses of vaccine).

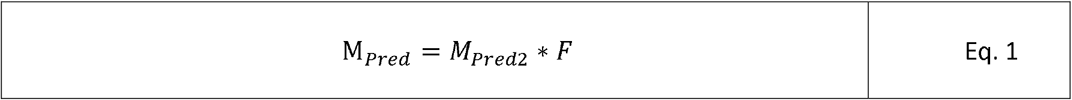

**Table 4.** Estimated rates of vaccine-attributable myocarditis/pericarditis cases post second dose of the primary series, by age subgroup, for 1 million fully vaccinated male individuals with Moderna CVmRNA.

| **Age Group** | **Moderna CVmRNA**<br>**Adjusted Rate per 1M Doses**<br>**Point Estimate and 95% CI** | **Data Source**<br>(DP = CBER BEST Data Partner) |
| --- | --- | --- |
| **18-25** | 127.8 [67.8, 241.2] | **DP1-4** |
| **26-35** | 31.8 [8.5, 118.9] | **DP1-4** |
| **36-45** | 23.1 [11.3, 47.2] | **DP1-4** |
| **46-55** | 12.5 [6.9, 22.8] | **DP1-4** |
| **56-64** | 9.6 [3.9, 23.3] | **DP1-4** |

*M*_*Pred2*_ is the meta-analysis estimated myocarditis/pericarditis case rates post dose 2. As presented previously^6^, these rates given in units of per 100k person-years were divided by 365 days and multiplied by 7 days (the risk window), which derives the total number of myocarditis/pericarditis cases within a 7-days risk window among the 100k people who completed their primary vaccination series. Next, we multiplied that number by 10 and derived the total number of cases among one million individuals who completed their primary vaccination series. Consequently, when converting from 100k person-years to one million vaccinated individuals’ risk post vaccination (in a 7-day risk window), the rates were multiplied by a factor *F* = (7*10)/365 as seen in Equation 1. We calculated the confidence intervals for the myocarditis cases with the chi-square method for Poisson distribution of rare events^12^. The number of myocarditis hospitalization (M_*H*_), ICU visits (M_*ICU*_), and deaths (M_*D*_) are fractions of predicted myocarditis/pericarditis cases (M_*Pred*_), such that M_*H*_ = M_*Pred*_* F_*H*,_ M_*ICU*_ = M_*Pred*_ * F_*ICU*_, and M_*D*_ = M_*Pred*_ * F_*D*_ where the scalars F_H_, F_ICU_, and F_D_ here are fractions of myocarditis cases that result in the more severe outcomes: hospitalizations, ICU visits, and deaths, respectively.

#### 2.3.2. Data and assumptions

##### 2.3.2.1. Myocarditis/pericarditis attributable to vaccine

We used myocarditis/pericarditis incidence data for male age groups derived from four BEST health claim databases (Table 4). Meta-analysis results are available for the incidence of myocarditis case rates within the risk window of 7 days by vaccine dose 1 and 2 separately. We limited this study to males 18-64 because the reported cases of myocarditis/pericarditis attributable to vaccine among female age groups and male and female age 65 years or above are very rare, leading to unstable estimates of case rates. Therefore, we are not able to provide reliable estimates of the risks for females in all age groups and individuals 65 years of age and above. Previous work on other mRNA vaccines has shown that the risks are lower for females and those over 65^15^.

##### 2.3.2.2. Myocarditis/pericarditis hospitalization and death rate

Vaccine Safety Datalink (VSD, a vaccine safety monitoring partnership between the CDC and 9 integrated healthcare organizations) data^13^ showed that 86% of myocarditis cases are hospitalized, and none required admission to the ICU for ages 18-29 years. The rate for hospitalization fell to 77% for ages 30-39 years. Since the age ranges in VSD differ from those in our analysis, we made the following adjustment for our age ranges. We used hospitalization rates of 86% for ages 18-25 years, 81.5% (the midpoint between ages 18-29 years and ages 30-39 years in VSD data) for ages 26-35 years, and 77% for ages 36 years and above in our analysis. The hospitalizations were mainly for monitoring patients’ condition as a precaution and no additional treatment is needed. Vaccine Safety Datalink (VSD) data show a median of a one-day stay during hospitalization^13^. No death confirmed to be caused by vaccine-attributable myocarditis/pericarditis has been identified. In this model, we used zero-death rate for vaccine attributable myocarditis/pericarditis as a model input.

## 3. Results

For Scenario 1 (base scenario), the model predicted that vaccination of 1 million males 18-25 years of age with two primary series doses of the vaccine would prevent 82,484 COVID-19 cases, 4,766 hospitalizations, 1,144 ICU admissions and 51 deaths due to COVID-19, while causing 128 vaccine-attributable myocarditis/pericarditis cases, 110 hospitalizations and no ICU admissions. No death due to vaccine attributed myocarditis/pericarditis is expected. These results represented the benefit-risk of the groups with the highest potential myocarditis/pericarditis risk under the current scenario (Omicron dominant, most recent peak incidence, and the mean rate of vaccine-attributable myocarditis/pericarditis case), and we considered that the benefits of vaccine clearly outweigh the risks. The results for all six model scenarios are summarized in Table 2, and we considered that the benefits of vaccine clearly outweigh the risks for each of the additional 5 scenarios as well. Scenarios 1, 2, and 3 represented the uncertainty in the case incidence (the week of December 25, 2021, average and the lowest incidence) in the future of the pandemic, Scenarios 1 and 4 represented the uncertainty in VE against the emerging variants (Omicron vs Delta), and Scenarios 1, 5, and 6 represented the uncertainty in the incidence rate of vaccine-attributable myocarditis/pericarditis.

## 4. Conclusions and Discussion

Based on available data, our results support the conclusion that the benefits of the Moderna vaccine clearly outweigh its risks for all the model scenarios for all males 18-64 years of age. Furthermore, based on consistent evidence that indicates a lower risk of vaccine-associated myocarditis in females of all ages and in males 65 years and older, it is reasonable to expect that the benefit-risk balance of vaccination with Moderna in these demographic groups would be even more favorable compared with males 18-64 years of age. Therefore, we concluded the benefits of the Moderna vaccine outweigh its risks for the overall target population.

Our modeling approach has a few limitations in the estimation of benefits. In this analysis, we conducted sensitivity analyses to test the impact of the COVID-19 incidence rates at the recent peak, average, and lowest incidence level of the pandemic. Nevertheless, given the uncertain trajectory of the pandemic, our constant COVID-19 incidence rate assumption creates high uncertainty on the benefit estimates. Furthermore, the percentage of hospitalizations resulting in ICU admission and the percentage of hospitalized patients who die are estimated based on cumulative rates of hospitalizations, ICU admissions, and deaths for each age group reported on COVID-NET from March 2020 to October 2021 (pre-Omicron period). The COVID-19 incidence rate might have changed since Omicron surged, but the more recent data were not yet available at the time of this analysis. The rate of ICU admission and death associated with Omicron may be lower compared to Delta, which may lead to overestimation of the benefits.

Also, estimated benefits of the vaccine would decrease if the vaccine became less effective against emerging SARS-CoV-2 variants. In this analysis, we evaluated the impact of different VEs for Omicron and Delta strains. However, there is uncertainty associated with future new variants or composition of the variants. Furthermore, the 30% VE for Omicron used in this analysis was obtained from a UK study. A similar US study with a smaller sample size showed an approximately 23% VE for Omicron within a 5-month period post second dose^14^. However, we do not expect that this difference in VE estimates would change our B-R conclusion. Another uncertainty for the model is vaccine protection durability. In this analysis, we assumed a 5-month protection period after completion of two primary doses of the vaccine overall. If vaccine-induced immunity significantly wanes within five months post second dose, potentially differentially for infection versus serious disease, that may reduce the benefit of the vaccine. Another limitation to the benefits is that we do not factor in the potential for a COVID-19 infection to impart additional protection. Antibody seroprevalence and its correlation to protection are not well understood yet. This could lead to an overestimation of the benefits of the vaccine. We assumed that the vaccine effectiveness against death would be equal to the vaccine effectiveness against COVID-19 hospitalizations in all scenarios. This could lead to underestimation of the benefits.

Our approach in estimating risks has limitations as well. First, neither the female group nor age 65 years or older of both sexes were included in this analysis due to the rarity of cases of myocarditis/pericarditis in these populations leading to an unstable estimate of case rate. However, the B-R for females and individuals 65 years of age or older is expected to be more favorable compared to male age 18-65 years, for whom clear favorable benefit-risk was demonstrated by this analysis. Second, there is uncertainty in the risk of myocarditis cases attributable to the vaccine. To estimate myocarditis/pericarditis risk attributable to the vaccine, health claims data were used, which have inherent limitations such as small sample sizes due to rare events among age groups. The reported cases in BEST had not been validated by a complete review of the patients’ medical charts, and therefore may be an overestimate. To address some of these limitations, the crude myocarditis rate in our model was adjusted using the myocarditis background rate in 2019 (measured by the BEST data partners), meta-analysis of four claims databases was conducted to increase the sample size, and a sensitivity analysis was conducted to test the impact on the benefit-risk of confidence interval of the estimated vaccine-attributable myocarditis/pericarditis case rate. Also, we use VSD data to calculate the rates of hospitalization and death associated with vaccine-attributable myocarditis/pericarditis. However, the actual rates we used for myocarditis came from the FDA’s BEST system, not VSD. The reason for this was that the VSD did not provide an age/sex specific myocarditis/pericarditis rate, and the young male group was our primary concern for the mRNA-based vaccine review. The BEST system, on the other hand, lacked information on hospitalizations and deaths caused by vaccine-attributable myocarditis and pericarditis. We did not estimate the proportions’ accuracy, which is especially important because the rates of myocarditis for VSD and BEST can differ.

Furthermore, some benefit-risk endpoints in our assessment are difficult to compare directly, for example, hospitalizations from COVID-19 and myocarditis hospitalizations. Our B-R assessment did not include potential long-term adverse effects due to either COVID-19 or myocarditis and second-order benefits and risks including potential impact on the public trust in COVID-19 vaccines. In this analysis, we did not investigate the benefits and risks for specific subpopulations, such as those with comorbidities, due to limited information for these populations. Health condition of individuals may remarkably impact the B-R profile and its evaluation.

Our model and analyses helped inform FDA’s licensure decision on Moderna’s vaccine. FDA considered the impact of COVID-19 pandemic on the public health, unavailability of treatment options, urgent need for a vaccine to prevent the disease and control the pandemic, together with available evidence and uncertainty associated with vaccine effectiveness and vaccine-attributable myocarditis/pericarditis risk. The FDA Review Committee agreed that the benefit/risk balance for the vaccine was favorable and supported approval for use of a 2-dose primary series in individuals 18 years of age and older^17^. The purpose for publication of this benefit-risk assessment is to increase the transparency of our regulatory action, communicating to the public that benefits of vaccination with the Moderna COVID-19 Vaccine clearly outweigh the risks even among the male adolescent population that is at a higher risk of myocarditis/pericarditis. This will hopefully increase public confidence in the vaccine and promote vaccination to fight COVID-19.

## Data Availability

All data produced in the present study are available upon reasonable request to the authors

## 5. Acknowledgements

We thank Christian Reed, Ph.D., Rituparna Moitra, Ph.D., and Rosser Matthews, Ph.D. for editing this manuscript. We thank Joyce Obidi, Ph.D., and Hui-Lee Wong, Ph.D. for helpful discussions. We thank, Diane Gubernot, Ph.D. and Marisabel Rodriguez Messan, Ph.D. for reviewing the manuscript. We thank FDA CBER OBPV data and analysis partners ACUMEN and OPTUM. Also, we thank the Centers for Disease Control and Prevention, particularly the Vaccine Task Force, for sharing initial benefit-risk assessment model and data on COVID-19 pandemic.

